# Models of COVID-19 vaccine prioritisation: a systematic literature search and narrative review

**DOI:** 10.1101/2021.06.21.21259104

**Authors:** Nuru Saadi, Y-Ling Chi, Srobana Ghosh, Rosalind M Eggo, Ciara McCarthy, Matthew Quaife, Jeanette Dawa, Mark Jit, Anna Vassall

## Abstract

**Background:** How best to prioritise COVID-19 vaccination within and between countries has been a public health and an ethical challenge for decision-makers globally. We systematically reviewed epidemiological and economic modelling evidence on population priority groups to minimise COVID-19 mortality, transmission and morbidity outcomes.

**Methods:** We searched the National Institute of Health iSearch COVID-19 Portfolio (a database of peer-reviewed and pre-print articles), Econlit, the Centre for Economic Policy Research and the National Bureau of Economic Research for mathematical modelling studies evaluating the impact of prioritising COVID-19 vaccination to population target groups. We narratively synthesised the main study conclusions on prioritisation and the conditions under which the conclusions changed.

**Findings:** The search identified 1820 studies. 36 studies met the inclusion criteria and were narratively synthesised. 83% of studies described outcomes in high-income countries. We found that for countries seeking to minimise deaths, prioritising vaccination of senior adults was the optimal strategy and for countries seeking to minimise cases the young were prioritised. There were several exceptions to the main conclusion, notably reductions in deaths could be increased, if groups at high risk of both transmission and death could be further identified. Findings were also sensitive to the level of vaccine coverage.

**Interpretation:** The evidence supports WHO SAGE recommendations on COVID-19 vaccine prioritisation. There is however an evidence gap on optimal prioritisation for low- and middle-income countries, studies that included an economic evaluation, and studies that explore prioritisation strategies if the aim is to reduce overall health burden including morbidity.

## Introduction

As of June 2021, over 1.5 billion vaccine doses have been administered, but vaccines are still in limited supply in the short to medium term in the vast majority of countries. [1] The question of which groups should be prioritised for vaccination within countries and between them has continued to present both a public health and an ethical challenge to decision makers. [2]

The World Health Organisation (WHO) Strategic Advisory Group of Experts on Immunization (SAGE) working group on COVID-19 vaccines has provided guidance to countries on the prioritisation of groups for vaccination while supply is limited. The guidance, based on the WHO SAGE values framework for the allocation and prioritization of COVID-19 vaccines, seeks to ensure equitable protection of human health across the globe, and in particular, among those experiencing the greatest risk and burden of COVID-19. [2, 3]

Epidemiological and economic models can provide an assessment of the potential health and broader societal impact of different prioritisation policies, and identify the optimal groups to prioritise for vaccination, given different public health objectives and scenarios. These results can be considered alongside other decision criteria to allocate vaccines both globally and within countries faced with a limited supply.

There was only a limit set of modelling results available to inform SAGE decision making at the end of 2020 (Figure 1), but in early 2021 the evidence base greatly expanded. The model results available at that time were largely limited to high-income and high-transmission settings such as the United States of America (USA) and United Kingdom (UK). Models specifically addressing low- and middle-income countries as well as low-transmission settings were not available.

**Figure 1.**
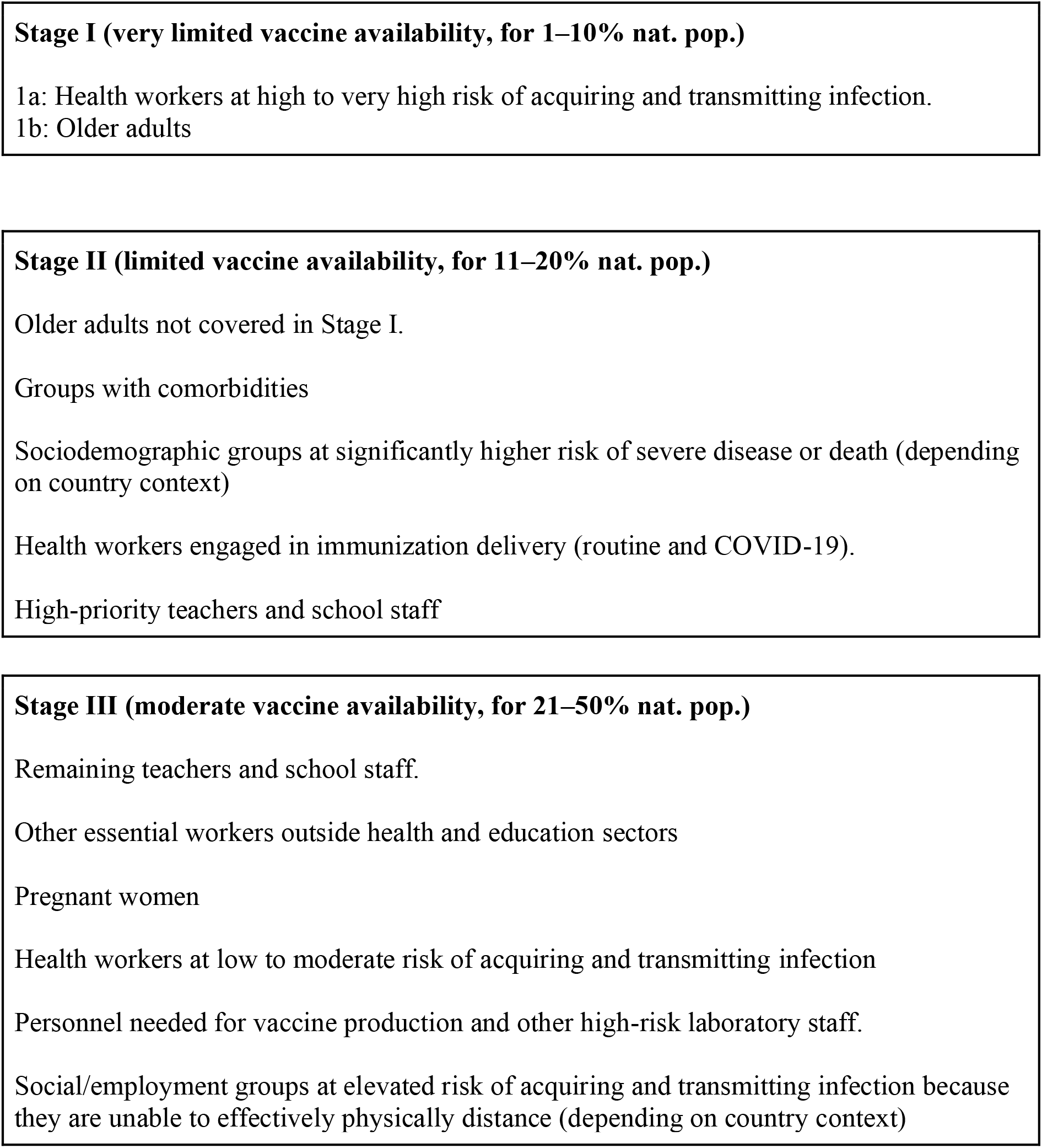
WHO SAGE vaccine prioritisation recommendations under different supply scenarios during community transmission (adapted from the WHO sage roadmap for prioritizing uses of COVID-19 vaccines in the context of limited supply)

We aimed to systematically review the epidemiological and economic modelling literature on population groups to prioritise for COVID-19 vaccination to minimise COVID-19 mortality, transmission and morbidity outcomes, in order to inform prioritisation policy at both the global and national levels. Our study is structured around the policy questions that SAGE considered in 2020. In summary, these questions examined exploring optimal allocation by age groups, occupational groups, groups with comorbidities and groups at higher risk of infection, considering the impact on deaths, cases, morbidity and economic outcomes. [3]

## Methods

### Search strategy and selection criteria

The systematic literature review was performed in line with PRISMA guidelines (Figure 2). [4] We searched the National Institute of Health (NIH) iSearch COVID-19 Portfolio on the 3^rd^ of March 2021 (a database which sources peer-reviewed articles from Pubmed and preprints from arXiv, bioRxiv, ChemRxiv, medRxiv, Preprints.org, Qeios, Research Square, and SSRN). We searched Econlit on the 3^rd^ of March 2021, using the advanced filters to include studies published between 2020 and 2021. To search these databases, we used a Boolean strategy to combine keywords such as “model*”, “vacc*”, “econom*”, “cost” and “COVID-19”. We contacted the Centre for Economic Policy Research in the United Kingdom and the National Bureau of Economic Research in the United States and received their full datasets of studies on the economics of COVID-19. (See Table S1 for the full search strategy and further details).

**Figure 2 -.**
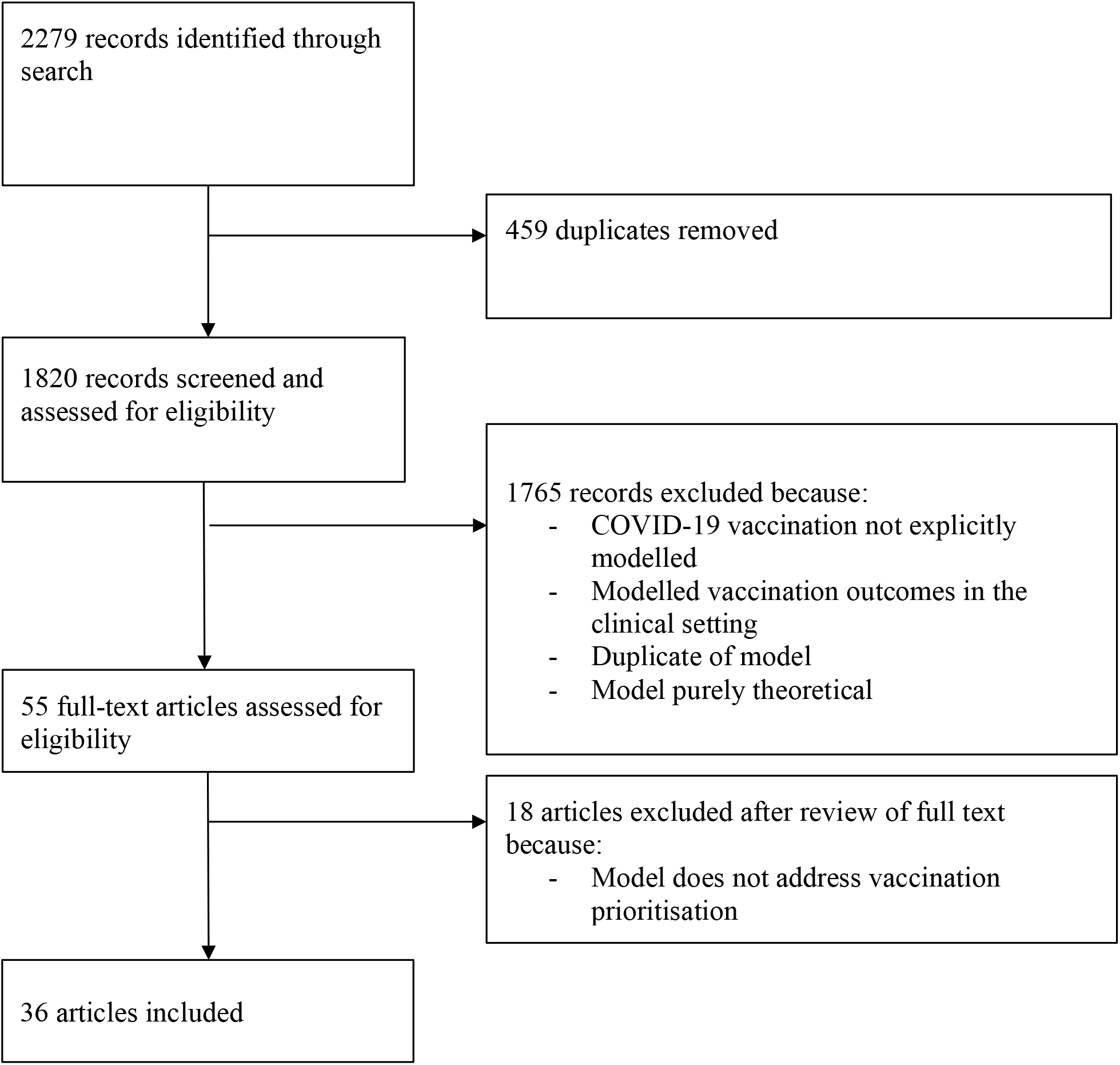
PRISMA diagram.

We included English language studies that used mathematical modelling to assess the impact of prioritising population target groups (either within or between countries) for COVID-19 vaccination on mortality, health (e.g. cases, quality adjusted life years), health care (e.g. hospitalisations) and/or economic (e.g. costs and cost-effectiveness) outcomes. We excluded studies which did not use a mathematical model to project the impact of COVID-19 vaccination, where none of the parameters were determined by empirical data (e.g. theoretical, non-empirical models), or which modelled vaccination outcomes within a clinical trial or a within-country small local setting, such as a care home, rather than nation or district wide allocation.

Two researchers independently screened titles and abstracts during the first round of screening. During the second round, three researchers independently screened titles/abstracts and full text articles. All studies were screened by at least two reviewers; where there were disagreements about inclusion these were resolved in discussion with a fourth researcher.

Three groups of two researchers each independently extracted the data from the included studies, with at least two groups reviewing each study. Discordant entries between the sets of extraction sheets were resolved by discussion between the groups.

Data were recorded in Microsoft Excel files summarising the policy objectives, outcomes, characteristics of the studies, the study conclusions and the conditions under which the conclusions changed i.e. sensitivity analyses (see supplementary materials to download the full extraction sheet).

Studies explored the optimal prioritisation strategy based on different policy objectives/ outcomes (deaths, cases etc.). We therefore extracted data and organised our results tables by the objective used. Some studies used two objectives to inform prioritisation (for example cases and deaths). In this instance we extracted and synthesised both sets of results. A full list of objectives of the included studies can be found in Table 1.

**Table 1 -.** Characteristics of studies.

| Characteristics | (n) |
| --- | --- |
| **Country** |  |
| HIC | 26 |
| MIC | 1 |
| LMIC | 3 |
| LIC | 0 |
| HIC, MIC & LIC | 2 |
| HIC & MIC | 1 |
| HIC, UMIC, LMIC & LIC | 1 |
| Theoretical | 1 |
| Not clear | 1 |
| **Model features** |  |
| Deterministic, compartmental | 23 |
| Stochastic, agent-based | 5 |
| Stochastic, compartmental | 4 |
| Deterministic, delay differential equation | 1 |
| Linear | 1 |
| Unclear | 2 |
| **Model structure** |  |
| SEIR | 11 |
| Expanded SEIR | 15 |
| Expanded SIR | 5 |
| SAPHIRE | 1 |
| Unclear natural history | 4 |
| **Contact matrix** |  |
| Age | 19 |
| Age & essential worker status | 1 |
| Age & day-specific | 1 |
| Age & location | 1 |
| Age, antibody status, major nationalities | 1 |
| Occupation, age, location & intensity of interaction | 1 |
| Social contact network | 3 |
| Vulnerable, front-line workers, non-vulnerable | 1 |
| Homogeneous mixing | 2 |
| Geographic mapping and socio-economic status | 1 |
| Jurisdiction contact rate (invariant with age) | 1 |
| Unclear | 4 |
| **Outcomes modelled** |  |
| Deaths | 29 |
| Cases | 23 |
| Hospitalisations | 11 |
| ICU admission | 5 |
| No of vx to avert one infection | 1 |
| Loss of economic benefits | 1 |
| Years of life lost | 3 |
| QALYs | 2 |
| DALYs | 1 |
| Cost-effectiveness ratio | 1 |
| Net present value of damages (VSL & DALYs) | 1 |
| Infection attack rate | 1 |
| Peak infections | 1 |
| Risk of new wave | 1 |
| Life-years gained | 1 |
| Total cost | 2 |
| Net monetary benefits | 1 |
| Effective reproductive number | 1 |
| Herd immunity | 1 |

We referred to the Economic and Social Research Council’s guidance on the conduct of narrative synthesis to aid data synthesis. [5] We therefore organised and grouped the textual results of the studies so that we could identify patterns within and between them. Synthesis was organised by the outcomes being explored. Due to wide variation between the studies in age group boundaries and other group classifications, extracted data from the study conclusions on vaccination priority groups were re-classified into broader population categories to aid synthesis. The population group categories considered were children/adolescents (ages 0-18), young adults (ages 19-40), middle-aged adults (ages 41-64), seniors (65+), groups with comorbidities, high social contact groups, essential workers, health workers and geographic regions.

Studies had different combinations of comparators, so we present results specifying the full range of comparator populations assessed. Study setting was categorised as High-Income (HIC), Upper Middle-Income (UMIC), Lower-Middle Income (LMIC), Low Income (LIC), multi-region or not specified, using World Bank classifications for 2021. [6] We extracted the modelling methods used, and reviewed assumptions and model structure in detail for the studies that did not align with the majority of conclusions to identify if this was based on an exceptional method (referred to as ‘exceptions’). We also report where sensitivity analyses generated results that were contrary to main findings of the study.

## Results

Our database search returned 2279 studies. After the removal of 459 duplicates, 1820 records were included in the title and abstract screening. After title and abstract screening, 55 studies remained for full text screening. After assessing the full text of the 55 studies for eligibility, 36 studies were included in the systematic review (see full extraction sheet Excel file in the supplementary materials and PRISMA flow diagram in Figure 2).

Study characteristics are summarised in Table 1. All the reviewed studies used models that captured transmission between individuals, with deterministic compartmental models being most common (23/36). However, agent-based models (5/36), stochastic compartmental models (4/36), a delay differential equation model (1/36), and a linear model (1/36) were also used. Studies most commonly used a SEIR (Susceptible, Exposed, Infected, Recovered) (11/36) or Expanded SEIR (15/36) natural history.

Most of them were set in a HIC (26/36); there were few single-country UMIC (1/36) and LMIC (3/36) studies. There were no single-country studies in a LIC setting. Only a few (4/36) looked at more than one country and two did not explicitly state the study setting. Most studies explored multiple policy objectives/outcomes regarding prioritisation: 28/36 investigated strategies to minimise deaths, 23/36 investigated minimisation of cases, 11/36 hospitalisations, 1/36 Quality Adjusted Life Years (QALYs), 1/36 Disability Adjusted Life Years (DALYs) and 3/36 Years of Life Lost (YLLs). Only 2/36 considered economic outcomes, such as financial or economic costs, in relation to prioritisation.

### Prioritisation to minimise deaths

Table 2 summarises the study conclusions highlighting the priority group and all the comparators included in each study (see methods section for how we defined population group categories). Most studies included seniors in the priority group. Thirteen studies recommended that seniors should be prioritised for vaccination to minimise deaths. [7] [8] [9] [10] [11] [12] [13] [14] [15] [16] [17] [18] [19] One study recommended prioritising seniors with comorbidities [20], and six studies recommended vaccinating seniors at the same time as another priority group (middle-aged adults, the highest social contact group, young and middle-aged adults who are in high contact with them, young adults with partial vaccine dose, and health workers). [21] [22] [23] [24] [25] [26]

**Table 2 -.** Studies aiming to minimise deaths (n=29)

| Conclusion on prioritisation to minimise deaths | No group prioritisation | No vaccination | Adolescents/c hildren | Young adults | Middle aged adults | Seniors | Comorbidities | According to size of populations (sub & national) | Social or work-related interaction | Forecasting of infected individuals | Other |
| --- | --- | --- | --- | --- | --- | --- | --- | --- | --- | --- | --- |
| **Seniors (20)<sup>1</sup>** | [14], [15], [26], [19] | [14], [15], [17], [20] | [7], [11], [12], [13], [16], [21], [22], [18], [23], [25], [19] | [7], [9], [10], [11], [12], [13], [15], [16], [17], [21], [22], [18], [23], [24], [25], [26] | [8], [9], [10], [11], [12], [13], [15], [16], [17], [21], [22], [18], [24], [23], [25], [19] | [11], [20], [18], [24], [19] | [11], [15], [20] | [18] | [7], [11], [14] |  | [25] |
| **Young adults (2)** |  |  |  | [24]* | [24]* | [33], [24]* |  |  |  |  |  |
| **High social contact groups (4)** | [31], [32], [29] |  | [31], [32], [23]* | [32], [23]* | [31], [32], [23]* | [31], [32] |  |  | [31], [32] | [29], [32] | [29] |
| **Essential workers (2)** |  |  | [27] | [27] |  | [27], [28] |  |  |  |  | [28] |
| **Other (3)** |  |  |  | [34] | [34] | [34] | [30] |  | [30] |  | [35] |

<sup>1</sup> Also includes: [21] & [22] seniors and middle aged adults, [26] seniors and health workers, [25] seniors at full dose and young adults at quarter dose, [28] young adult and middle aged essential workers, [24] seniors & young and middle aged group in high contact with them, [23] high social contact group & seniors. **Other:** [34] school-going youth, [35] by serology testing, [30] non-vulnerable group. \*Appears twice

Ten studies did not find that prioritising the groups at highest risk of mortality (seniors or people living with comorbidities) minimised deaths. [27] [28] [29] [30] [31] [32] [33] [24] [23] [34] These ‘exceptional’ studies instead found that prioritising groups with a higher risk of infection would lead to fewer deaths; in other words, that the indirect protection from lower transmission outweighs the benefits from direct protection from vaccines for those at the highest risk of mortality. The group at high risk of infection were defined as those with: higher contact rates e.g. synthetic population with 3-10 times the average number of contacts of the age group 30-39 [23]; individuals with an expansive social network [31] [32]; and individuals with essential worker status. [27] [28] In addition, two studies defined young adults as the group with the highest social interactions and therefore at higher risk of infection. [29] [33] One study examined vaccination of individuals that had high levels of interaction with seniors. [24]

One of the ten exceptional studies concluded that the non-vulnerable group should be prioritised for vaccination compared to the group with comorbidities. [30] In this study the authors state they assumed that “the time required to vaccinate the vulnerable group is identical to that of the much larger non-vulnerable group”, even though the non-vulnerable group was much larger. Finally, one study recommended prioritising vaccination through the use of serological testing to achieve the greatest reduction in COVID-19 related deaths. [35]

### Prioritisation to minimise COVID-19 cases

Table 3 summarises the study conclusions. The largest proportion of the selected studies (N=23) investigated optimal vaccine prioritisation strategies to minimise COVID-19 cases. Of these, six studies recommended young and middle aged adults, [10] [16] [12] [22] [15] [17] one young adults, [33] and two young adults and children. [21] [26] One study recommended young people at the same time as another priority group (seniors at full vaccine dose and young adults at partial dose). [25]

**Table 3 -.** Studies aiming to minimise cases (n=23)

| Conclusion on optimal prioritisation to minimise cases | No group prioritisation | No vaccination | Adolescents/children | Young adults | Middle aged adults | Seniors | Comorbidities | According to size of populations (sub & national) | Allocation by geographic disease burden | Social or work-related interaction | Other |
| --- | --- | --- | --- | --- | --- | --- | --- | --- | --- | --- | --- |
| **Young adults (11)<sup>2</sup>** | [12], [15], [26] | [15], [17] | [16], [12], [22], [25] | [16], [15], [25] | [16], [22], [15], [21], [28], [25] | [10], [16], [12], [22], [15], [17], [21], [26], [28], [33] | [15] |  |  | [26], [28] | [28], [25] |
| **High social contact group (4)** | [31], [32], [37] |  | [31], [32] | [32] | [31], [32] | [31], [32], [37] |  |  |  | [31], [32], [36] | [32], [37] |
| **Essential workers (3)** | [14] | [14] | [27] | [27] | [28]* | [14], [27], [28]* |  |  |  | [14], [27], [28]* | [28]* |
| **Seniors (3)** |  |  | [13], [18], [25]* | [13], [18], [25]* | [13], [18], [25]* | [18] |  | [18] | [18] |  | [25]* |
| **Other (4)** | [40] | [38] | [38] | [38] | [38], [40] | [38], [40] | [40] | [39] | [39] |  | [35], [38], [40], [39] |

<sup>2</sup> Also includes young and middle-aged adults, young adults and children. Other: [40] all age groups equally, [35] & [38] by serology testing, [39] area of low disease burden. \*Appears twice

Seven studies recommended vaccination prioritisation based on social or occupational interactions compared to age group prioritisation. [28] [27] [31] [32] [36] [14] [37] Of these, three studies recommended prioritising essential workers to minimise cases [28] [27] [14] and four studies recommended prioritising high social contact adults compared to other age groups. [31] [32] [37] [36]

Two studies recommended prioritising vaccination using serological testing to prioritise antibody-negative individuals compared to not using serological testing, [35] [38] and a further study recommended that the geographic area with lower disease burden should be prioritised for vaccination compared to the geographic area with high disease burden. [39]

There were a few studies concluding differently to the majority recommendations on minimising cases. Three studies found that scenarios targeting seniors [13] [18] [25] led to the highest reduction in cases. However, two of those studies did not have a comparator that modelled those strategies comparted to more socially interactive populations. [18] [25] Chhetri et al found very small differences between scenarios, and the conclusion was not reported in the results section. [13]

### Prioritising other outcomes

Studies investigating strategies to minimise hospitalisations from COVID-19 tended to reach similar conclusions to studies investigating deaths (N=11). Seven studies recommended prioritising senior adults [8] [15] [16] [41] [37], senior and middle-aged adults [21] or seniors and the high social contact group [23] for vaccination compared to other age and occupational groups. 4 studies concluded differently from the majority of the hospitalisation outcome studies. [31] [32] [38] [40] Two recommended prioritising the high social contact group compared to prioritising senior adults. [31] [32] One study recommended prioritising vaccination by serological testing compared to no serological testing. [38] One study recommended giving equal priority to all age and risk groups compared to a targeted age-based prioritisation. [40]

A few studies investigated the optimal vaccination strategy when maximising QALYs, DALYs or YLLs. One study modelled the vaccination prioritisation strategy to minimise QALY losses. [11] The authors concluded that the most effective strategy to minimise QALY losses is to prioritise senior adults for vaccination compared to other age groups, groups with comorbidities and no group prioritisation. Three studies investigated within-country vaccine prioritisation strategies for minimising YLLs. [12] [28] [22] Two studies recommended prioritising seniors for vaccination to minimise YLLs [12][28] and the other recommended prioritising middle aged adults and seniors. [22] One study modelled the impact of COVID-19 vaccination on DALYs. [19] The authors found that the amount of DALYs averted under a base vaccination strategy which prioritised seniors was stable to a scenario where everyone over 15 years old is vaccinated. [19]

One study considered the cost-effectiveness of COVID-19 vaccination. [19] The authors found that the strategy of prioritising seniors for vaccination was similarly cost-effective to vaccinating all individuals over 15 years old. [19]

One study investigated prioritisation strategies for optimising the incremental net monetary benefit (iNMB) of vaccination, i.e. the net economic gain from vaccination including both costs saved and monetised health gains. [40] The authors concluded that giving equal priority to all age and risk groups was most optimal compared to prioritising seniors, high risk individuals, and both seniors and high risk individuals when vaccine effectiveness was only moderate (40%) and coverage was low (20%). Conversely, when vaccine effectiveness was high (80%) and coverage was moderate (50%), vaccinating high risk individuals resulted in the highest iNMB.

### Prioritisation by setting

Three of the included articles were single-country studies modelling LMIC settings. [14] [17] [19] One study modelled an UMIC setting. [33] These studies reached the same conclusions as the HIC studies apart from one: those minimising deaths recommended prioritising seniors, while those minimising cases recommended prioritising high transmission groups. The exception was one study from Thailand on minimising cases which recommended prioritising high transmission groups to minimise deaths. [33]

Additionally, there were three multi-country studies which modelled LMIC settings [8] [9] [12] and one modelling an UMIC setting. [23] The conclusions for these studies were in line with the majority conclusions for deaths and cases (except for one study which recommended prioritising both the high social contact group and seniors to minimise deaths). [23] See Table S5 for a summary of the studies modelling UMIC and LMIC settings.

One study also considered prioritisation between countries, in addition to within countries. This study made recommendations on global vaccine allocation strategies to optimise different health objectives. [9] The authors concluded that the strategy to minimise deaths was to allocate doses equitably across all income settings relative to population size, and then to prioritise vaccination of seniors within countries. This performed better than allocating vaccines to countries based on their respective senior population sizes, giving preferential allocation to HICs, giving preferential allocation to LICs and LMICs, or allocating doses in proportion to population plus providing a set number of extra doses to HIC and UMICs. [9] When YLLs were used as an optimisation outcome measure, LMIC settings received the most doses.

### Factors that influence prioritisation strategy

35 out of 36 (97%) studies included a sensitivity analysis. Of these, 14 studies reported a sensitivity analysis that led to a potential change in the recommended prioritisation strategy. Whilst there were a wide range of parameters tested in the uncertainty analysis there were only a few that consistently drove a change in prioritisation. The most common parameters that influenced prioritisation all related to vaccine coverage (level of vaccine supply, vaccine coverage, and speed of vaccine rollout).

Six studies reported that the trade-off between direct and indirect protection is sensitive to the proportion of people vaccinated. [12] [9] [21] [22] [15] [32] These papers stated that when vaccine supply is very low vaccination has a minimal impact on interrupting transmission, so more deaths can be prevented to vaccinate groups at risk of severe disease (e.g., key workers, seniors and clinical risk groups). However, as supply increases, this opens up the possibility of interrupting transmission, which can change prioritisation to the young or those with many contacts. If there is very high vaccine supply, seniors are again favoured for prioritisation if aiming to reduce deaths, as there is sufficient coverage to achieve both direct protection of the most vulnerable and indirect protection from key transmitters.

Several studies tested different values of vaccine efficacy, with most reporting before full results of vaccine trials became available starting in late 2020. Generally, variations in vaccine efficacy did not appear to change prioritisation unless efficacy was significantly lower in older rather than younger populations. However, a number of studies assumed that vaccines had similar levels of efficacy against severe disease, infection or transmission. Where vaccines were more efficacious against severe disease strategies, the priority was to vaccinate highest transmitters.

## Discussion

We find that for countries seeking to minimise deaths, the current evidence base supports prioritising vaccination of senior adults (65+) as the optimal strategy, unless there are exceptional cases where specific non-age-related high-risk groups or very highly networked individuals can be identified and prioritised. The difference in deaths averted can be large between depending on the strategies, for example in one study, a symptom-blocking vaccine with 50% uptake prioritising seniors and high-risk groups averted 17000 more deaths than an unprioritized campaign. [20] For countries seeking to minimise cases, the evidence supports prioritising young age groups and essential workers. The evidence base examining the optimal strategy to improve health in general is too limited to draw any firm conclusions.

While in principle prioritising highly socially connected groups may be optimal to reduce mortality, it could prove difficult in practice to identify these groups, especially when their definition and identification are only vaguely defined. [31] [32] [24] [23] Chen et al. suggest those population groups could be identified through contact tracing, although recognising the limitations of such an approach in resource constrained settings. [23] Santini recommends prioritising younger people with many connections to vulnerable people. [24] Buckner et al. find that prioritising essential workers (based on occupation) could lead to fewer deaths in the context of strong non-pharmaceutical interventions. [28] As no studies included the feasibility and costs of identifying and delivering vaccines to highly-connected groups, it is unclear whether prioritisation to groups that are not age or occupation-based is possible or cost-effective.

Luangasanatip et al. examined how vaccines may be prioritised in a low-incidence setting (Thailand). [33] finding that prioritising younger age groups would lead to greater reductions in deaths. However, this was the only study set in a low incidence setting, so more research may be required to validate this finding across settings and modelling approaches. The small number of studies set in low- and middle-income countries, and the lack of evidence on the cost-effectiveness of reaching different population also limits our findings in resource constrained settings.

Although we found that most of the studies modelling these settings were in line with the conclusions from studies set in HIC for minimising deaths and cases, context may impact results, especially when very limited supply is considered. The studies based in LMIC settings assumed a higher level of vaccine supply available and level of coverage achieved than has been observed in most LMICs (see Table S5). Moreover, many LMICs have not yet established senior adult vaccination programmes, so targeting by age may be challenging and costly. Further research is urgently required to model the effect of different levels of supply on prioritisation in lower income settings.

There was only one study modelling inter-country allocation of vaccines, despite the political importance of this issue. [42] That one study found that doses should be allocated equally by population size if minimising deaths, and allocated preferentially to low- and middle-income countries if minimising life years lost. Since this differs drastically from the current allocation of vaccine doses globally, it points to the need for further investigation and action. [9]

Our review found that the optimal prioritisation strategy to pursue depends on the public health objective(s) of the decision-maker, with different conclusions depending on whether the objective is reducing cases or reducing mortality. [31] [32] The trade-offs between different objectives are a challenging ethical issue for decision-makers.

The WHO SAGE values framework for the allocation and prioritization of vaccines proposed 6 principles as the ethical basis of decisions on vaccine prioritisation: the promotion of human well-being, equal respect, global equity, national equity, reciprocity and legitimacy. [2] Within the framework, reducing disease burden overall (and not purely the number of deaths) is a key consideration to promote human wellbeing. However, only one study considered integrated burden of disease outcomes such as QALYs that combine both morbidity and mortality in relation to prioritisation [11], one study considered DALYs. [19] This evidence gap may be particularly limiting in settings with a younger population, such as in many low- and middle-income countries, where overall mortality may be a smaller proportion of the overall COVID-19-related burden morbidity compared to high-income countries.

Only one study considered within-country equity (such as prioritising populations that have suffered disproportionately from COVID-19 because of their socioeconomic status). [37] We also consider the few economic studies, such as economic evaluations, to represent a research gap. The choice of one vaccine strategy over another in the studies evaluated often only took into account the net health gain, yet the choice of the most appropriate vaccination strategy should take into account health benefits, costs and the willingness to pay threshold - which varies in each setting. [43]

Our findings are limited by several methodological issues. By limiting our search to English language studies, we may have missed relevant studies, particularly in low- and middle-income countries. Furthermore, much of this literature is pre-print studies which are not peer-reviewed, so the quality of the evidence presented here should be viewed with caution. Finally, to highlight key findings across all studies, we categorised the reviewed studies according to the broad public health objectives that they aimed to fulfil. However, this categorisation may have obscured some nuances within studies, such as where there were variations in study conclusions grouped under the same category.

## Conclusion

The findings of this systematic literature review have provided empirical evidence for the prioritisation of senior adults for vaccination to minimise COVID-19 deaths and young people to minimise COVID-19 transmission. However, there remain critical gaps in the evidence around strategies that reduce overall health outcomes, consider the costs of different prioritisation strategies and for low- and middle-income settings. The research gaps identified can help to guide the direction of further research on vaccination prioritisation as the pandemic continues to evolve.

## Supporting information

Data extraction sheet

## Data Availability

The extracted data from the studies referred to in the manuscript is available in the supplementary material.

## Bibliography

[1] WHO, “WHO Coronavirus (COVID-19) Dashboard,” [Online]. Available: covid19.WHO.int. [Accessed 8 June 2021..

[2] WHO, “WHO SAGE values framework for the allocation and prioritization of COVID-19 vaccination,” World Health Organization, 2020.

[3] WHO, “WHO Strategic Advisory Group of Experts (SAGE) on Immunization Working Group on COVID-19 Vaccines: Prioritized Infectious Disease and Economic Modelling Questions,” World Health Organization, 2020.

[4] A. Liberati, D. G. Altman, J. Tetzlaff, C. Mulrow, P. C. Gøtzsche, J. P. A. Ioannidis, M. Clarke, P. J. Devereaux, J. Kleijnen and D. Moher, “The PRISMA statement for reporting systematic reviews and meta-analyses of studies that evaluate healthcare interventions: explanation and elaboration,” BMJ, vol. 339, no. b2700, 2009.

[5] J. Popay, H. Roberts, A. Sowden, M. Petticrew, L. Arai, M. Rodgers, N. Britten, K. Roen and S. Duffy, “Guidance on the Conduct of Narrative Synthesis in Systematic Reviews,” ESRC, 2006.

[6] T. W. Bank, “World Bank Country and Lending Groups,” 2021. [Online]. Available: https://datahelpdesk.worldbank.org/knowledgebase/articles/906519-world-bank-country-and-lending-groups. [Accessed 8 June 2021].

[7] P. Jentsch, M. Anand and C. T. Bauch, “Prioritising COVID-19 vaccination in changing social and epidemiological landscapes,” 27 September 2020. [Online]. Available: https://www.medrxiv.org/content/10.1101/2020.09.25.20201889v2. [Accessed 9 June 2021].

[8] M. T. Meehan, D. G. Cocks, J. M. Caldwell, J. M. Trauer, A. I. Adekunle, R. R. Ragonnet and E. S. McBryde, “Age-targeted dose allocation can halve COVID-19 vaccine requirements,” 2 December 2020. [Online]. Available: https://www.medrxiv.org/content/10.1101/2020.10.08.20208108v2.full. [Accessed June 8 2021].

[9] B. A. Hogan, P. Winskill, O. J. Watson, P. G. Walker, C. Whittaker, M. Baguelin, D. Haw, A. Løchen, K. A. M. Gaythorpe, I. C. C.-1. R. Team, F. Muhib, P. Smith, K. Hauck and Fer, “Report 33: Modelling the allocation and impact of a COVID-19 vaccine,” Imperial College COVID-19 response team, 2020.

[10] X. Chen, M. Li, D. Simchi-Levi and T. Zhao, “Allocation of COVID-19 Vaccines Under Limited Supply,” 26 September 2020. [Online]. Available: https://www.medrxiv.org/content/10.1101/2020.08.23.20179820v2. [Accessed June 8 2021].

[11] S. Moore, E. M. Hill, L. Dyson, M. J. Tildesley and M. J. Keeling, “Modelling optimal vaccination strategy for SARS-CoV-2 in the UK,” 24 September 2020. [Online]. Available: https://www.medrxiv.org/content/10.1101/2020.09.22.20194183v2. [Accessed 8 June 2021].

[12] K. M. Bubar, K. Reinholt, S. M. Kissler, M. Lipsitch, S. Cobey, Y. H. Grad and D. B. Larremore, “Model-informed COVID-19 vaccine prioritization strategies by age and serostatus,” 8 January 2021. [Online]. Available: https://www.medrxiv.org/content/10.1101/2020.09.08.20190629v3.full.pdf. [Accessed 8 June 2021].

[13] B. Chhetri, D. K. K. Vamsi, S. Balasubramanian and C. B. Sanjeevic, “Optimal Vaccination and Treatment Strategies in Reduction of COVID-19 Burden,” 19 February 2021. [Online]. Available: https://arxiv.org/abs/2102.09802. [Accessed 8 June 2021].

[14] J. M. A. Minoza, V. P. Bongolan and J. F. Rayo, “COVID-19 Agent-Based Model with Multi-objective Optimization for Vaccine Distribution,” 27 January 2021. [Online]. Available: https://arxiv.org/abs/2101.11400. [Accessed 8 June 2021].

[15] N. Hoertel, M. Blachier, F. Limosin, M. Sánchez-Rico, C. Blanco, M. Olfson, S. Luchini, M. Schwarzinger and H. Leleu, “Optimizing SARS-CoV-2 vaccination strategies in France: Results from a stochastic agent-based model,” 20 January 2021. [Online]. Available: https://www.medrxiv.org/content/10.1101/2021.01.17.21249970v1. [Accessed 8 June 2021].

[16] T. N.-A. Tran, N. Wikle, J. Albert, H. Inam, E. Strong, K. Brinda, S. M. Leighow, F. Yang, S. Hossain, J. R. Pritchard, P. Chan, W. P. Hanage, E. M. Hanks and. M. F. Boni, “Optimal SARS-CoV-2 vaccine allocation using real-time seroprevalence estimates in Rhode Island and Massachusetts,” 15 January 2020. [Online]. Available: https://www.medrxiv.org/content/10.1101/2021.01.12.21249694v1. [Accessed 8 June 2021].

[17] B. H. Foy, B. Wahl, K. Mehta, A. Shet, G. I. Menon and C. Britto, “Comparing COVID-19 vaccine allocation strategies in India: a mathematical modelling study,” 24 November 2020. [Online]. Available: https://www.medrxiv.org/content/10.1101/2020.11.22.20236091v1.full. [Accessed 8 June 2021].

[18] D. Bertsimas, J. Ivanhoe, A. Jacquillat, M. Li, A. Previero, O. S. Lami and H. T. Bouardi, “Optimizing Vaccine Allocation to Combat the COVID-19 Pandemic,” 18 November 2020. [Online]. Available: https://www.medrxiv.org/content/10.1101/2020.11.17.20233213v1.full. [Accessed 8 June 2021].

[19] C. A. B. Pearson, F. Bozzani, S. R. Procter, N. G. Davies, M. Huda, H. T. Jensen, M. Keogh-Brown, M. Khalid, S. Sweeney, S. Torres-Rueda, C. C.-1. W. G. and C. C.-1. W. G., “Health impact and cost-effectiveness of COVID-19 vaccination in Sindh Province, Pakistan,” 25 February 2021. [Online]. Available: https://www.medrxiv.org/content/10.1101/2021.02.24.21252338v1. [Accessed 9 June 2021].

[20] X. Wang, Z. Du, K. E. Johnson, S. J. Fox, M. Lachmann, J. S. McLellan and L. A. Meyers, “The impacts of COVID-19 vaccine timing, number of doses, and risk prioritization on mortality in the US,” 20 January 2020. [Online]. Available: https://www.medrxiv.org/content/10.1101/2021.01.18.21250071v1. [Accessed 8 June 2021].

[21] L. Matrajt, J. Eaton, T. Leung and E. R. Brown, “Vaccine optimization for COVID-19, who to vaccinate first?,” 16 August 2020. [Online]. Available: https://www.medrxiv.org/content/10.1101/2020.08.14.20175257v1. [Accessed 8 June 2021].

[22] E. Shim, “Optimal Allocation of the Limited COVID-19 Vaccine Supply in South Korea,” J. Clin. Med, vol. 10, no. 591, 2021.

[23] M. Moret, T. R. Filho, J. Mendes, T. Murari, A. N. Filho, A. Cordeiro, W. Ramalho, F. Scorza and A.-C. Almeida, “WHO vaccination protocol can be improved to save more lives,” 18 Jan 2021. [Online]. Available: https://www.researchsquare.com/article/rs-148826/v1. [Accessed 8 June 2021].

[24] S. Santini, “Covid-19 vaccination strategies with limited resources --a model based on social network graphs,” 11 October 2020. [Online]. Available: https://arxiv.org/abs/2010.05312. [Accessed 8 June 2021].

[25] P. Hunziker, “Impact of personalized-dose vaccination in Covid-19 with a limited vaccine supply in a 100 day period in the U.S.A.,” 16 February 2021. [Online]. Available: https://www.medrxiv.org/content/10.1101/2021.01.30.21250834v4. [Accessed 8 June 2021].

[26] C. R. Macintyre, V. Costantino and M. Trent, “Modelling of COVID-19 vaccination strategies and herd immunity, in scenarios of limited and full vaccine supply in NSW, Australia,” 19 December 2020. [Online]. Available: https://www.medrxiv.org/content/10.1101/2020.12.15.20248278v2. [Accessed 8 June 2021].

[27] A. Babus, S. Das and S. Lee, “The Optimal Allocation of Covid-19 Vaccines,” 3 December 2020. [Online]. Available: https://www.medrxiv.org/content/10.1101/2020.07.22.20160143v2. [Accessed 8 June 2021].

[28] J. H. Buckner, G. Chowell and M. R. Springborn, “Dynamic Prioritization of COVID-19 Vaccines When Social Distancing is Limited for Essential Workers,” 06 October 2020. [Online]. Available: https://www.medrxiv.org/content/10.1101/2020.09.22.20199174v4. [Accessed 8 June 2021].

[29] J. Rodriguez, M. Paton and J. M. Acuna, “COVID-19 vaccine prioritisation to the groups with the most interactions can substantially reduce total fatalities,” 30 November 2020. [Online]. Available: https://www.medrxiv.org/content/10.1101/2020.10.12.20211094v2. [Accessed 8 June 2021].

[30] M. B. Bonsall, C. Huntingford and T. Rawson, “Optimal time to return to normality: parallel use of COVID-19 vaccines and circuit breakers,” 3 February 2021. [Online]. Available: https://www.medrxiv.org/content/10.1101/2021.02.01.21250877v1. [Accessed 8 June 2021].

[31] J. Chen, S. Hoops, A. Marathe, H. Mortveit, B. Lewis, S. Venkatramanan, A. Haddadan, P. Bhattacharya, A. Adiga, A. Vullikanti, A. Srinivasan, M. L. Wilson, G. Ehrlich and M. Fenster, “Prioritizing allocation of COVID-19 vaccines based on social contacts increases vaccination effectiveness,” 16 February 2021. [Online]. Available: https://www.medrxiv.org/content/10.1101/2021.02.04.21251012v2.full.pdf. [Accessed 8 June 2021].

[32] B. Goldenbogen, S. O. Adler, O. Bodeit, J. A. Wodke, X. Escalera-Fanjul, A. Korman, M. Krantz, L. Bonn, R. Morán-Torres, J. E. Haffner, M. Karnetzki, I. Maintz, L. Mallis, H. Prawitz and Segelit, “Optimality in COVID-19 vaccination strategies determined by heterogeneity in human-human interaction networks,” 18 December 2020. [Online]. Available: https://www.medrxiv.org/content/10.1101/2020.12.16.20248301v1. [Accessed 8 June 2021].

[33] N. Luangasanatip, W. Pan-Ngum, J. Prawjaeng, S. Saralamba, L. White, R. Aguas, H. Clapham, C. Painter, W. Yi, W. Isaranuwatchai and Y. Teerawattananon, “Optimal vaccine strategy to control COVID-19 pandemic in middle-income countries: Modelling case study of Thailand,” 24 February 2021. [Online]. Available: https://www.researchsquare.com/article/rs-270635/v1. [Accessed 8 June 2021].

[34] A. D. Visscher, B. Sutton and T. Sutton, “Second-wave dynamics of COVID-19: Impact of behavioral changes, immunity loss, new strains, and vaccination,” 22 February 2021. [Online]. Available: https://www.researchsquare.com/article/rs-195879/v1. [Accessed 8 June 2021].

[35] A. B. Fujimoto, I. Yildirim and P. Keskinocak, “Significance of SARS-CoV-2 Specific Antibody Testing during COVID-19 Vaccine Allocation,” 1 February 2021. [Online]. Available: https://www.medrxiv.org/content/10.1101/2021.01.28.21250721v1. [Accessed 8 June 2021].

[36] B. Shayak and M. M. Sharma, “COVID-19 Spreading Dynamics with Vaccination - Allocation Strategy, Return to Normalcy and Vaccine Hesitancy,” 11 December 2020. [Online]. Available: https://www.medrxiv.org/content/10.1101/2020.12.10.20247049v1. [Accessed 9 June 2021].

[37] S. C. Bruningk, J. Klatt, M. Stange, A. Mari, M. Brunner, T.-C. Roloff, H. M. Seth-Smith, M. Schweitzer, K. Leuzinger, K. K. Søgaard, D. A. Torres, A. Gensch and A.-. Schlotterbeck, “Determinants of SARS-CoV-2 transmission to guide vac2 cination strategy in a city,” 17 December 2020. [Online]. Available: https://www.medrxiv.org/content/10.1101/2020.12.15.20248130v2. [Accessed 9 June 2021].

[38] H. H. Ayoub, H. Chemaitell, M. Makhoul, Z. A. Kanaani, E. A. Kuwari, A. A. Butt, P. Coyle, A. Jeremijenko, A. H. Kaleeckal, A. N. Latif, R. Mohammad, M. G. A. Kuwari and H. E. Romaihi, “Epidemiological impact of prioritizing SARS-CoV-2 vaccination by antibody status: Mathematical modeling analyses,” 12 January 2021. [Online]. Available: https://www.medrxiv.org/content/10.1101/2021.01.10.21249382v1. [Accessed 9 June 2021].

[39] F. M. Castonguay, J. C. Blackwood, E. Howerton, K. Shea, C. Sims and J. N. Sanchirico, “Spatial Allocation of Scarce Vaccine and Antivirals for COVID-19,” 19 January 2021. [Online]. Available: https://www.medrxiv.org/content/10.1101/2020.12.18.20248439v3. [Accessed 9 June 2021].

[40] E. Kirwin, E. Rafferty, K. Harback, J. Round and C. McCabe, “A Net Benefit Approach for the Optimal Allocation of a COVID-19 Vaccine,” 2 December 2020. [Online]. Available: https://www.medrxiv.org/content/10.1101/2020.11.30.20240986v1.full. [Accessed 9 June 2021].

[41] S. Guerstein, V. Romeo-Aznar, M. Dekel, O. Miron, N. Davidovitch, R. Puzis and S. Pilosof, “Optimal strategies for combining vaccine prioritization and social distancing to reduce hospitalizations and mitigate COVID19 progression,” 22 December 2020. [Online]. Available: https://www.medrxiv.org/content/10.1101/2020.12.22.20248622v1.full. [Accessed 8 June 2021].

[42] O. J. Wouters, K. C. Shadlen, M. Salcher-Konrad, A. J. Pollard, H. J. Larson, Y. Teerawattananon and M. Jit, “Challenges in ensuring global access to COVID-19 vaccines: production, affordability, allocation, and deployment,” The Lancet, vol. 397, no. 10278, pp. P1023–1934, 2021.

