## Supplementary tables for "Models of COVID-19 vaccine prioritisation: a systematic literature search and narrative review"

### Supplementary materials

**Table S1: Search strategy**

| Database | Search terms | Dates |
|---|---|---|
| National Institute of Health iSearch COVID-19 portfolio<br><br>(peer-reviewed articles from Pubmed and Preprints from arXiv, bioRxiv, ChemRxiv, medRxiv, Preprints.org, Qeios, Research Square, and SSRN) | [title and abstract]<br><br>(1) model* AND vacc* AND (math* OR computation* OR simulat* OR optim*)<br><br>(2) vaccin* AND (econom* OR cost) | 03/03/2021 |
| CEPR & NBER | All economics papers on COVID (hand search) | 03/03/2021 |
| Econlit | [Title and abstract]<br><br>Cost AND (covid-19 or coronavirus or 2019-ncov or sars-cov-2 or cov-19) | Entire database searched on 03/03/2021 (filtered 2020-2021) |

**Table S2 - Exceptions: death outcome studies**

| Author | Conclusions |
|---|---|
| Luangasanatip et al | Prioritise young adults |
| Chen (J) et al | Prioritise high social contact group |
| Bonsall et al | Prioritise the non-vulnerable group |
| Goldenbogen et al | Prioritise high social contact group |
| Moret et al | Prioritise high social contact group + seniors |
| Visscher et al | Prioritise children |
| Santini | Prioritise both seniors and the middle-age and young adults who have high-contact with them |
| Rodriguez et al | Prioritise high social contact group |
| Buckner et al | Prioritise young adult and middle-age essential workers, then elderly |

| | |
|---|---|
| Babus et al | Prioritise middle-aged adult workers |

**Table S3 - Exceptions: cases outcome studies**

| **Author** | **Conclusions** |
|---|---|
| Chhetri et al | Prioritise seniors |
| Kirwin et al | Give equal priority to all age and risk groups |
| Bertsimas et al | Prioritise geographic areas with higher projected cases and the seniors within those areas |
| Hunziker et al | Prioritise seniors at full dose and young adults at quarter-dose in parallel |

**Table S4: Exceptional studies by level of coverage**

| Coverage level by end of vx campaign (% pop) | Deaths exceptions | Cases exceptions |
|---|---|---|
| 50-100 | 4 | 3 |
| 30-50 | | 1 |
| 1-30 | 1 | |
| N/A | 4 | |
| | 9 | 4 |

#### Table S5 - Summary of UMIC and LMIC studies

| F Author | Setting | Conclusion deaths | Conclusion cases | Supply and coverage |
|---|---|---|---|---|
| Minoza | Philippines | Seniors | Mobile workforce (non-medical) | 500k vx to cover 1/6 of pop (unclear timing) |
| Foy | India | Seniors | Young and middle-aged adults | 6 months-4 yrs to cover 100% of target pop |
| Pearson | Pakistan | Seniors | | 20% of pop phase 1. 4000 vx pd initially then increase in quarters over 1 year (base) |
| Meehan | 179 countries (detailed results for India, China & UK) | Seniors | | Depends on country and scenarios: up to 100% (timing not specified) |
| Bubar | Belgium; US; India; Spain; Brazil; Zimbabwe; South Africa; China | Seniors | Young and middle-aged adults | 1-50% total pop supply (0.2% pd until supply exhausted) |
| Hogan | Global (HIC/UMIC/LMIC/LIC) | Seniors | | Individuals vaccinated at a constant rate over a 1 month in country. 2 billion vx global constraint |
| Luangasanatip | Thailand | Young adults | Young adults | 9M vx in 2 months |
| Moret | Brazil and Portugal | High social contact group + seniors | | 250M vx for Brazil & 20M for Portugal available in one year |

### Table S6 - Sensitivity analysis tables

| **Studies** | **Outcomes** | **Original priority** | **Priority changed in sensitivity analysis** | **Sensitivity analysis that changes the priority** |
|---|---|---|---|---|
| Ayoub | Cases | Serology based | | |
| Babus, A | Cases | Middle aged adult workers | Yes | Increasing vaccine supply would decrease the age of the youngest eligible recipients. |
| | Deaths | | | |
| Bertsimas, D | Cases | E/V | | |
| | Deaths | E/V | | |
| Bonsall, M | Deaths | Y/HC | | |
| Brüningk, S | Cases | HC (low SES) | | |
| Bubar, K | Cases | Y/HC | | |
| | Deaths | E/V | Yes | Change priority to 20-49 if:<br><br>VE low in older adults, R0 = 1.15 and VE >80% transmission blocking, R0=1.15 and rollout speed 0.2% per day or vaccine for 25% of the population, leaky vaccines (prioritise children). |
| Buckner, J | Cases | Y/HC | | |
| | Deaths | Y/HC | | |
| Castonguay, F | Cases | Area with the lower disease burden | | |
| Chen, J | Cases | Y/HC | | |

| | | | | |
|---|---|---|---|---|
| | Deaths | Y/HC | | |
| Chen, X | Cases | E/V | Yes | For dynamic policies, the older groups should be vaccinated at early days and then switch to younger. |
| | Deaths | E/V | | |
| Chhetri, B | Cases | E/V | | |
| | Deaths | E/V | | |
| Foy, B | Deaths | E/V | | |
| | Cases | Y/HC | | |
| Fujimoto, A | Cases | Serology based | | |
| | Deaths | | | |
| Goldenbogen, B | Cases | Y/HC | Yes | For high vaccination levels the strategy to vaccinate the most interactive individuals first is most effective for all three objectives. There is a trade-off between different strategies for low levels of vaccination. |
| | Deaths | Y/HC | | |
| Grundel, S | | Y/HC | | |
| Guerstein, S | | E/V | | |
| Hogan, A | Deaths | E/V | Yes | For limited supply (<20%) target elderly/vulnerable. For supply >20% switch to targeting key transmitters (potentially include children) for indirect protection. |
| Hoertel, N | Cases | Y/HC | Yes | With lower supply prioritise the older population. |
| | Deaths | E/V | | |
| Hunziker, P | Cases<br>Deaths | Prioritise seniors at 100% and | | |

| | | | | |
|---|---|---|---|---|
| | | younger at 25% dose | | |
| Jentsch, P | Deaths | E/V | Yes | If seropositivity is high (particularly if roll out is delayed) or VE is lower in vulnerable groups, prioritise to interrupt transmission. |
| Kirwin, E | Cases | Equal priority to all groups | Yes | Rank ordering of different prioritisation options varied greatly by prioritisation criteria, with different vaccine effectiveness and coverage, and by concurrently implemented policies. |
| Luangasanatip, N | Cases | Y/HC | Yes | VE reduction in severity 70-90% and reduced susceptibility 0%, prioritise vulnerable. |
| | Deaths | Y/HC | | |
| Macintyre, C R | Cases | Y/HC | | |
| | Deaths | E/V | | |
| Matrajit, L | Cases | Y/HC | Yes | Vaccine coverage for approx. half the population with VE>60%, prioritise high transmission. If more vaccine becomes available, prioritise the vulnerable again. (Depends on pre-existing immunity). |
| | Deaths | E/V | | |
| Meehan, M | Cases | Y/HC | | |
| | Deaths | E/V | | |
| Minoza, J | Cases | Mobile work force | | |
| | Deaths | E/V | | |
| Moore, S | Deaths | E/V | Yes | When R>1 and there is a race to herd-immunity to prevent further rise a rapid vaccine deployed untargeted campaign is far more successful than a slow but optimally targeted one. If vaccine efficacy is significantly reduced in the elderly (<20%) then other orderings become more effective. |

| | | | | |
|---|---|---|---|---|
| Moret, M | Deaths | E/HC | | |
| Pearson, C | Deaths | E/V | | |
| Rodriguez, J | Deaths | HC | | |
| Shayak, B | Cases | Y/HC | | |
| Shim, E | Cases | Y/HC | Yes | Supply >80% incidence-minimizing strategies lead to a broader vaccination strategy for those aged 10–69 years. |
| | Deaths | E/V | | |
| Tran, T | Cases | Y/HC | | |
| | Deaths | E/V | | |
| Visscher, A | Deaths | Children | Yes | It is likely that small changes in the assumptions underlying the calculations will reverse these conclusions. It is more reasonable to conclude that, as long as an adequate level of vaccination is reached, it is of less importance who is prioritized. |
| Wang, X | Deaths | E/V | Yes | Prioritisation has limited benefit under low (<50%) uptake. |

Abbreviations: E; elderly, HC; high contacts, SES; socio-economic status, V; vulnerable, VE; vaccine efficacy, Y; younger population
